# Comparative analysis of the first wave of the COVID-19 pandemic in South Korea, Italy, Spain, France, Germany, the United Kingdom, the USA and the New-York state

**DOI:** 10.1101/2020.11.20.20235689

**Authors:** Luis Alvarez

## Abstract

We use an exponential growth model to analyze the first wave of the COVID-19 pandemic in South Korea, Italy, Spain, France, Germany, the United Kingdom, the USA and the New-York state. This model uses the number of officially reported patients tested positive and deaths to estimate an infected hindcast of the cumulative number of patients who later tested positive or who later die. For each region, an epidemic timeline is established, obtaining a precise knowledge of the chronology of the main epidemiological events during the full course of the first wave. It includes, in particular, the time that the virus has been in free circulation before the impact of the social distancing measures were observable. The results of the study suggest that among the analyzed regions, only South Korea and Germany possessed, at the beginning of the epidemic, a testing capacity that allowed to correctly follow the evolution of the epidemic. Anticipation in taking measures in these two countries caused the virus to spend less time in free circulation than in the rest of the regions. The analysis of the growth rates in the different regions suggests that the exponential growth rate of the cumulative number of infected, when the virus is in free circulation, is around 0.250737. In addition, we also study the ability of the model to properly forecast the epidemic spread at the beginning of the epidemic outbreak when very little data and information about the coronavirus were available. In the case of France, we obtain a reasonable estimate of the peak of the new cases of patients tested positive 9 days in advance and only 7 days after the implementation of a strict lockdown.

## 1 Introduction

In this work we analyze the first wave of the COVID-19 pandemic in South Korea, Italy, Spain, France, Germany, the United Kingdom, the USA and the New-York state using the number of officially reported patients tested positive and deaths. We use the exponential growth model introduced in [1] to estimate the infected hindcast who later tested positive or who later die. To obtain an approximation of the observable data we use the temporal distribution of the incubation period (in the case of patients tested positive) or the temporal distribution from infection to death (in the case of deaths). Let us denote by *y*(*t*) the cumulative number of infected. We can obtain an approximation, *N*(*t*), of the cumulative number, *D*(*t*), of patients tested positive (or deaths) using the formula:

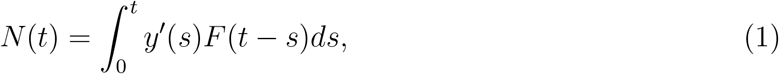

where *F*(*t*) is the cumulative distribution of the incubation time (or the temporal distribution from infection to death). As we will explain in more details later, we assume that *y*(*t*) follows an exponential growth with a time-varying growth rate *r*(*t*) which depends on some parameters that are adjusted so that *N*(*t*) is as close to *D*(*t*) as possible. *N*(*t*) can be interpreted as a smooth version of *D*(*t*) that allows the calculation of epidemiological events such as the peak of the number of daily cases. As illustrated in Fig. 1 we identify 3 phases in the development of the first epidemic wave: the first phase begins at the epidemic outbreak and ends after passing the peak of daily cases when the number of daily cases is already descending. The second phase is the consolidation of the descent and the third phase is the stabilization around a baseline. The number of days of each phase is manually fixed in the algorithm to get the best approximation between *N*(*t*) and *D*(*t*). Throughout these 3 phases we study the following epidemiological events on the observed data: The date of the epidemic outbreak, the date when a lockdown was implemented (if that event occurs). The date that social distancing measures begin to take effect on the observed data (denoted by **FTM** in the figure), the date that the peak is reached, the date that the number of daily cases is divided by two, and finally the date when the number of daily cases stabilizes (denoted by **EFW** in the figure).

**Figure 1:**
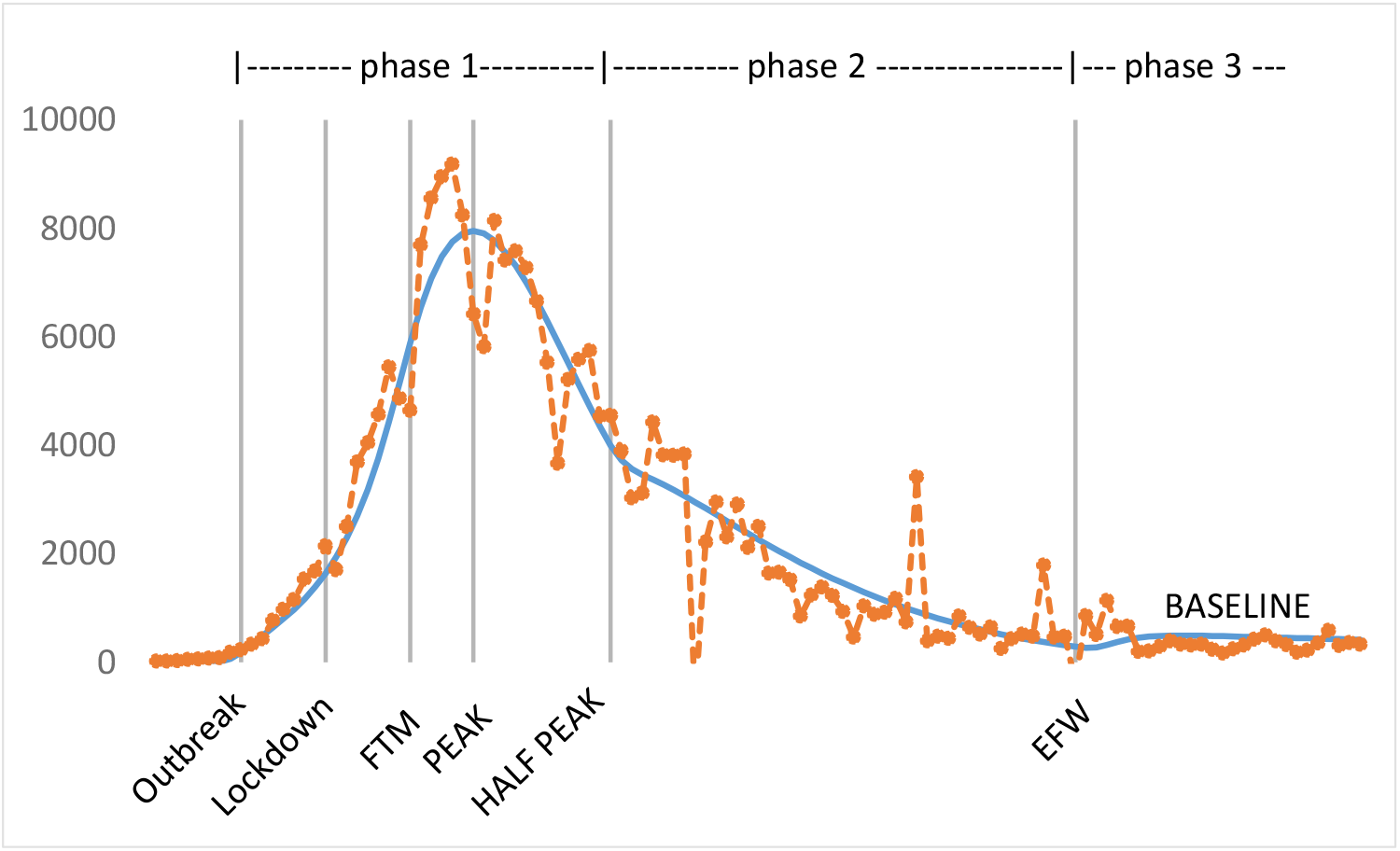
In the case of Spain, we show, in the early stages of the epidemic spread: the recorded daily number of patients tested positive *D*′(*t*) (dashed line) and its approximation using *N*′(*t*) (solid line). We also illustrate the 3 phases that we consider in the development of the first epidemic wave and the epidemiological events described in the text.

We use this methodology to study the chronology of the main epidemiological events during the full course of the first wave for the evolution of the number of patients tested positive and deaths. In both cases we estimate an infected hindcast, *y*(*t*). Analyzing these infected hindcast we can estimate the exponential growth rate at the beginning of the epidemic spread. In fact, we have experimentally observed that this exponential growth rate is very similar for the infected hindcast in the case of patients tested positive and in the case of death. This means that, at the beginning of the epidemic spread, the way in which the number of infected who later test positive grows and the way in which the number of infected who later die grows are very similar.

Moreover, the comparison of the shift between the same epidemiological event for patients tested positive and deaths provides valuable information about the testing capacity in the region. For instance, the better the testing capacity, the greater the shift between the dates of the observed initial outbreak of patients tested positive and the one of deaths.

In addition, we also study the ability of the model proposed in [1] to properly forecast the epidemic spread at the beginning of the epidemic outbreak when very little data and information about the coronavirus were available. In this case, the goal is not to study the full course of the first wave, the goal is to estimate the model using the recorded data up to a given day and to study the forecast error when predicting the date of the daily peak of the number of new cases or the number of daily new cases 3 weeks after the peak. In the case of France, for patients tested positive, we obtain a reasonable forecast of the peak 9 days in advance and only 7 days after the implementation of a strict lockdown.

The rest of the paper is organized in the following way: in section 2 we present the exponential growth model that we use and its relation with the SIR models. In section 3, we present a comparative analysis of the obtained chronology of the main epidemiological events for the different regions. In section 4, we present, in the case of France, a study of the ability of the model for forecasting in the early stages of the epidemic spread and finally section 4 concludes.

## 2 The exponential growth model

We assume that *y*(*t*), the infected hindcast, grows at an exponential rate (that we name *a*), during a period of time *t*_0_. We thus have *y*′(*t*) = *ay*(*t*). Then, after strict social distancing measures are implemented the exponential rate *r*(*t*) (such that *y*′(*t*) = *r*(*t*)*y*(*t*)) decreases until it attains the value 0 at time *t*_1_. We model this behavior using the following empirical time-varying exponential rate:

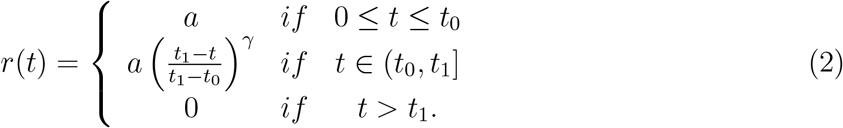

where *a, γ, t*_0_ and *t*_1_ *> t*_0_ are positive numbers which determine the parameters of the model. *r*(*t*) is a continuous non-increasing function which is well adapted to track the evolution of the epidemic in its early stages when the implementation of the social distancing measures produces a decrease in the exponential growth rate of *y*(*t*). We point out that the time *t*_1_ where *r*(*t*) becomes zero can be beyond the interval time where we apply the model, that is, we are not forcing *r*(*t*) to become zero in the interval time of interest.

As we will see later, this model can be considered as a plausible approximation of the basic SIR model at the beginning of the epidemic, when the number of recovered can be neglected and the measures of social distancing require the introduction of empirical functions to model the variation in the exponential growth rate in the SIR model. The underlying basic differential equation *y*′(*t*) = *r*(*t*)*y*(*t*) can be solved explicitly, and in the case of *r*(*t*) given by (2) the solution is:

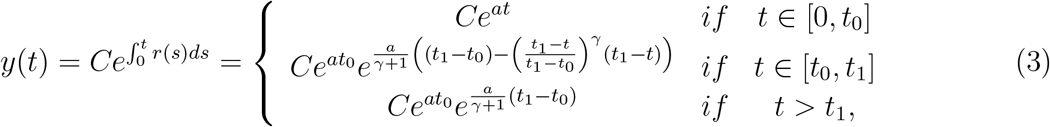

we notice that *C* and *t*_0_ are closely related because if we change *t*_0_ by *t*_0_ − *T, C* by *Ce*^*aT*^ and *t*_1_ by *t*_1_ − *T*, the solution *y*(*t*) does not change. Therefore *t*_0_ is an “abstract” time and we can not say that the coronavirus has been actually in free circulation for *t*_0_ days. What we can say is that the coronavirus has been in free circulation until *y*(*t*) reaches the value 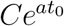 but, from the above formula, we can not decide the actual starting time of the epidemic outbreak.

The asymptotic state of the number of contaminated subjects is

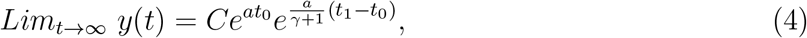

and it is attained at *t*_1_. Therefore the impact of the social distancing measures is determined by the value:

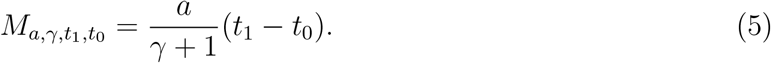

The smaller this value, the more effective the social distancing interventions. We notice that the peak in the new daily contaminated patients is obtained when *y*′(*t*) changes sign which corresponds to *y*″(*t*) = 0. Using a straightforward computation we obtain that the peak is attained at

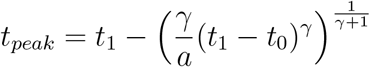

From *y*(*t*) and the cumulative distribution *F*(*t*) we compute an approximation, *N*(*t*), of the observable data using equation (1). In the results presented in this paper we use as distribution for the incubation period the one proposed in Lauer et al. in [6], given by a log-normal distribution of parameters *µ* = 1.621 and *σ* = 0.418. For the distribution from infection to death we use the one proposed in [1] given by a mixture of two log-normal distributions of parameters *µ* = 1.621, *σ* = 0.418) and *µ* = 2.351375257, *σ* = 0.6011434688.

### 2.1 Relation with the SIR model

The basic SIR model separates the population in three compartments: *S*(*t*) (the number of susceptible), *I*(*t*) (the number of infectious), and *R*(*t*) (the number of recovered). It should be mentioned that in this model the number of deaths is not considered. In that sense, we can assume that R(t) is the sum of recovered and deceased. Each member of the population typically progresses from susceptible to infectious to recovered. The basic SIR model to estimate *S*(*t*), *I*(*t*) and *R*(*t*) is the following system of ordinary differential equations:

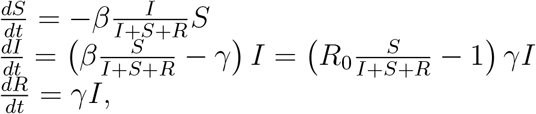

where *β* and *γ* are parameters which depend on the particular disease. 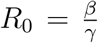, named the reproductive number, is one of the key parameters in transmission models and it represents the number of secondary infections that arise from a typical primary case in a completely susceptible population. Notice that *S*(*t*), the number of susceptible subjects, is a decreasing function. When the ratio between *S*(*t*) and the total population satisfies

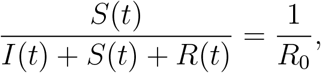

we obtain 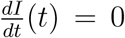. Hence the peak of infected subjects is attained, and from that time, the number of infected subjects starts decreasing. Notice that the larger *R*_0_, the larger the time required to attain the infection peak. We observe that in our model, in the evolution of contaminated patients, *y*(*t*), we include the infected and recovered subjects, so *y*(*t*) = *I*(*t*)+*R*(*t*) and then using the SIR model we obtain that

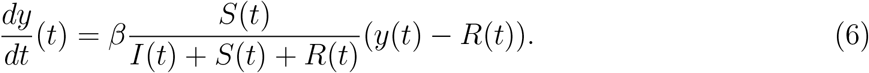

The SIR model with constant *γ* and *β* and the conclusions about the peak of infected subjects make sense only if the virus propagates freely across time, but everything changes if we impose social distancing measures to the population. A natural way to include human interventions in the SIR model is to replace *β* by a time dependent function *β*(*t*). This strategy has been used by different authors in different contexts using extended versions of the SIR models. For instance in [3], the authors propose the following exponential type function:

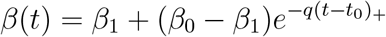

another exponential type function has been introduced in [7]:

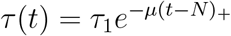

In [2] the following rational function is proposed:

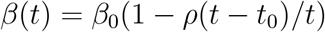

In [4] the author proposes the function

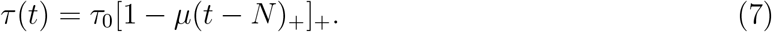

We observe that this is a particular case of the function (2) defining *r*(*t*) where *a* = *τ*_0_, *µ* = 1*/*(*t*_1_− *t*_0_), *N* = *t*_0_ and *γ* = 1. The only difference of this function with *r*(*t*) is that in *r*(*t*) we add the power *γ* to modulate the way the exponential growth rate decreases.

In this work we assume that social distancing interventions govern the evolution of contaminated subjects rather than the SIR dynamic and we replace equation (6) by

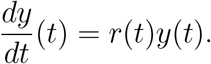

Therefore we include in the term *r*(*t*) the impact of the human interventions, the influence of the ratio between *S*(*t*) and the total population and the influence of *R*(*t*). The latter makes sense if we are at the beginning of the pandemic (so *R*(*t*) ≈ 0) or if we assume that *R*(*t*) is proportional to *y*(*t*). By focusing just on the number of contaminated subjects we reduce the complexity of the problem and we avoid to deal with the balance between infected, exposed and recovered patients which is very difficult to estimate properly due to the lack of accuracy in the data we can manage about the number of infected subjects.

It can be objected that the model we use for *r*(*t*) defined by (2) is empirical, but to our knowledge, so far, the functions that have been proposed to include time-varying exponential growth *β*(*t*) in the SIR models are also empirical and their purpose is to try to fit empirically the SIR model solution to observed data when social distancing measures are implemented.

### 2.2 An extended model to track the full course of the first epidemic wave

The exponential growth given by equation (2) is very simple and it is useful to compute an estimation of *y*(*t*) after an strict lockdown is implemented, this estimation covers from the epidemic outbreak until a certain time after the daily peak. However if we want to go further and to approximate the evolution for a longer time we need to extend the model to have more flexibility in order to fit the epidemic spread. In fact, the above basic model can be easily extended in the following way: let 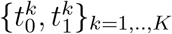 be 2 increasing sequences of real numbers satisfying 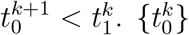 represent times where a change is expected in the evolution trend of the epidemic. Then the exponential growth (2) can be extended in the following way:

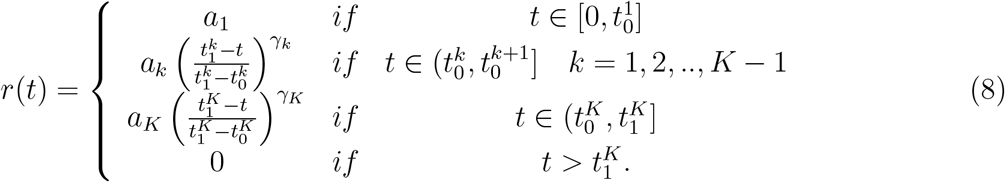

Notice that *r*(*t*) can be discontinuous at 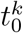 because a relaxation of social distancing measures will definitely produce an abrupt modification in the exponential growth of the epidemic. We point out that the function *r*(*t*) is always decreasing except at the possible points of discontinuity 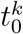. In particular, the model is not well adapted to scenarios where the growth rate can grow continuously, such as a second epidemic wave.

In this work, to study the full-course of the first epidemic wave, we use a basic model and 2 extended models (that is *K* = 3). These three models address the three phases of the first epidemic wave explained in the introduction, that is: the first phase begins at the epidemic outbreak and ends after passing the peak of daily cases when the number of daily cases is already descending, the second phase is the consolidation of the descent and the third phase is the stabilization around a baseline.

All the experiments we have performed can be reproduced using the algorithm implemented in the online interface www.ctim.es/covid19. The algorithm fits the parameters of the models by minimizing a quadratic error between the observed data *D*(*t*) and its approximation *N*(*t*). The main parameters of the algorithm are the number of days, *N*_*k*_, assigned manually to each of the described phases and the option to fix the value of the efficiency of the social distancing measures given by 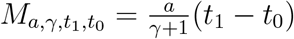. For more details about the algorithm see [1].

We used the dataset of the evolution of patients tested positive or deaths for the different countries from the web page https://www.ecdc.europa.eu/en/publications-data/download-todays-data-geographic-distribution-covid-19-cases-worldwide. All the experiments presented can be reproduced using the online interface in https://ipolcore.ipol.im/demo/clientApp/demo.html?id=301.

## 3. Comparative analysis of the full course of the epidemic during the first wave in the different regions

We are going to use the exponential model explained above to try to better understand the evolution of the first epidemic wave in South Korea, Italy, Spain, France, the United Kingdom, the USA and the New York state. Among other things, we are going to estimate the number of days that the virus has been circulating freely before the effect of the social distancing measures take effect, as well as the exponential growth rate of infected in this phase of free circulation. We are going to study the effect of a strict lockdown implemented in the early stages of the epidemic spread, the time it takes to reach the peak of daily cases and the time it takes to divide by two the number of cases reached at the peak. We also compare the results obtained for patients tested positive and deaths, which provide interesting information on the testing capacity of these regions at the beginning of the epidemic spread.

For each region we manually chose the parameters of the algorithm in the online interface www.ctim.es/covid19 to get the best fit between the daily registered data and the model prediction given by *N*′(*t*) (see (1)). In Fig. 3, 4, 5 and 6 we illustrate the results obtained and the model parameters for each region. For each region, the value of the efficiency 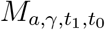only appears if it is manually fixed in the algorithm.

**Figure 2:**
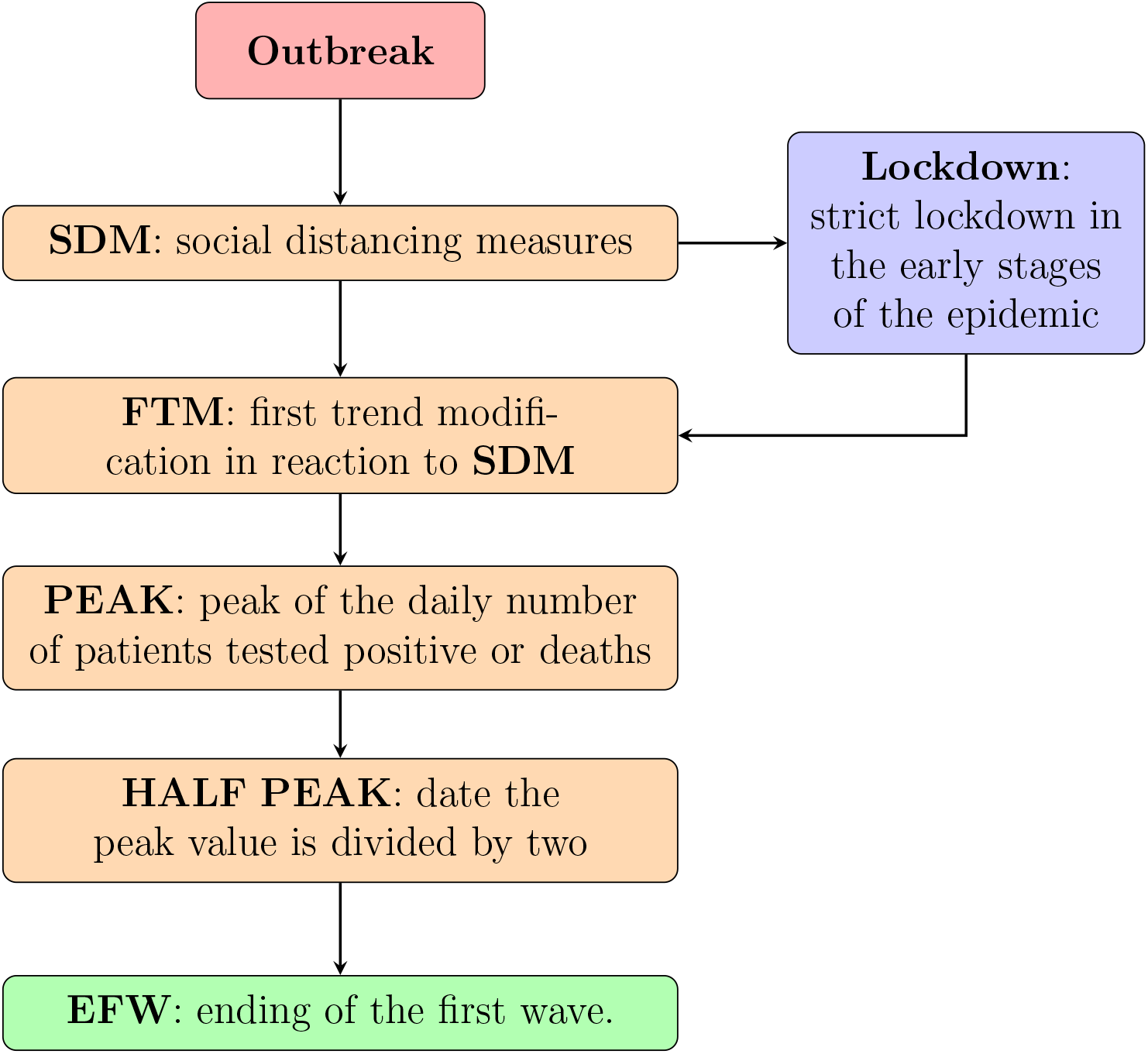
Timeline of events we consider in the first wave of the COVID-19 epidemic spread.

**Figure 3:**
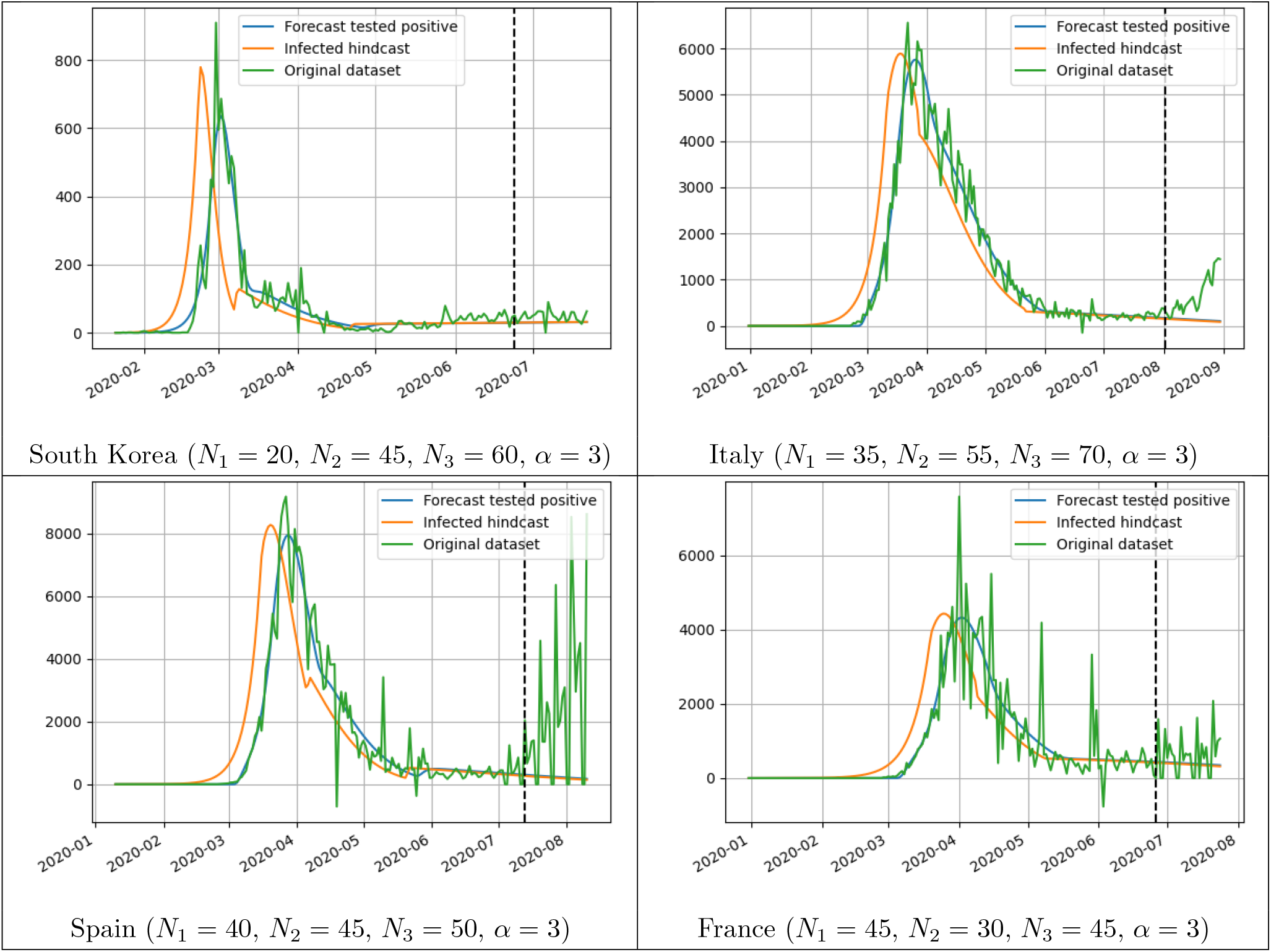
Extended model for patients tested positive applied to South Korea, Italy, Spain and France. Below the plot of each country we show the parameters of the model manually chosen to get the best fit between the daily registered data and the model prediction.

**Figure 4:**
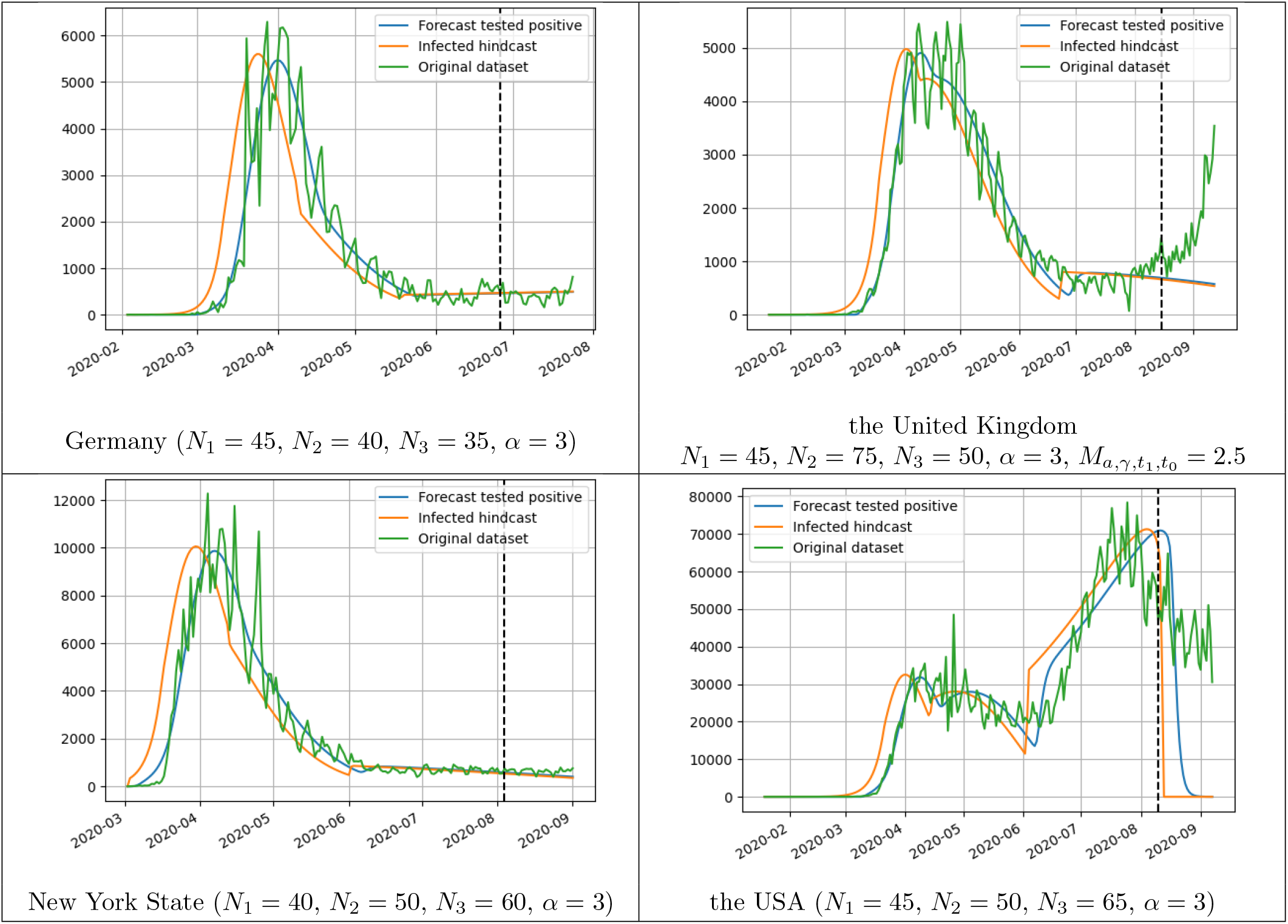
Extended model for patients tested positive applied to Germany, the United Kingdom, New York State and the USA. Below the plot of each country we show the parameters of the model manually chosen to get the best fit between the daily registered data and the model prediction. The value of 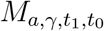(see (5)) only appears if it is manually fixed in the algorithm.

**Figure 5:**
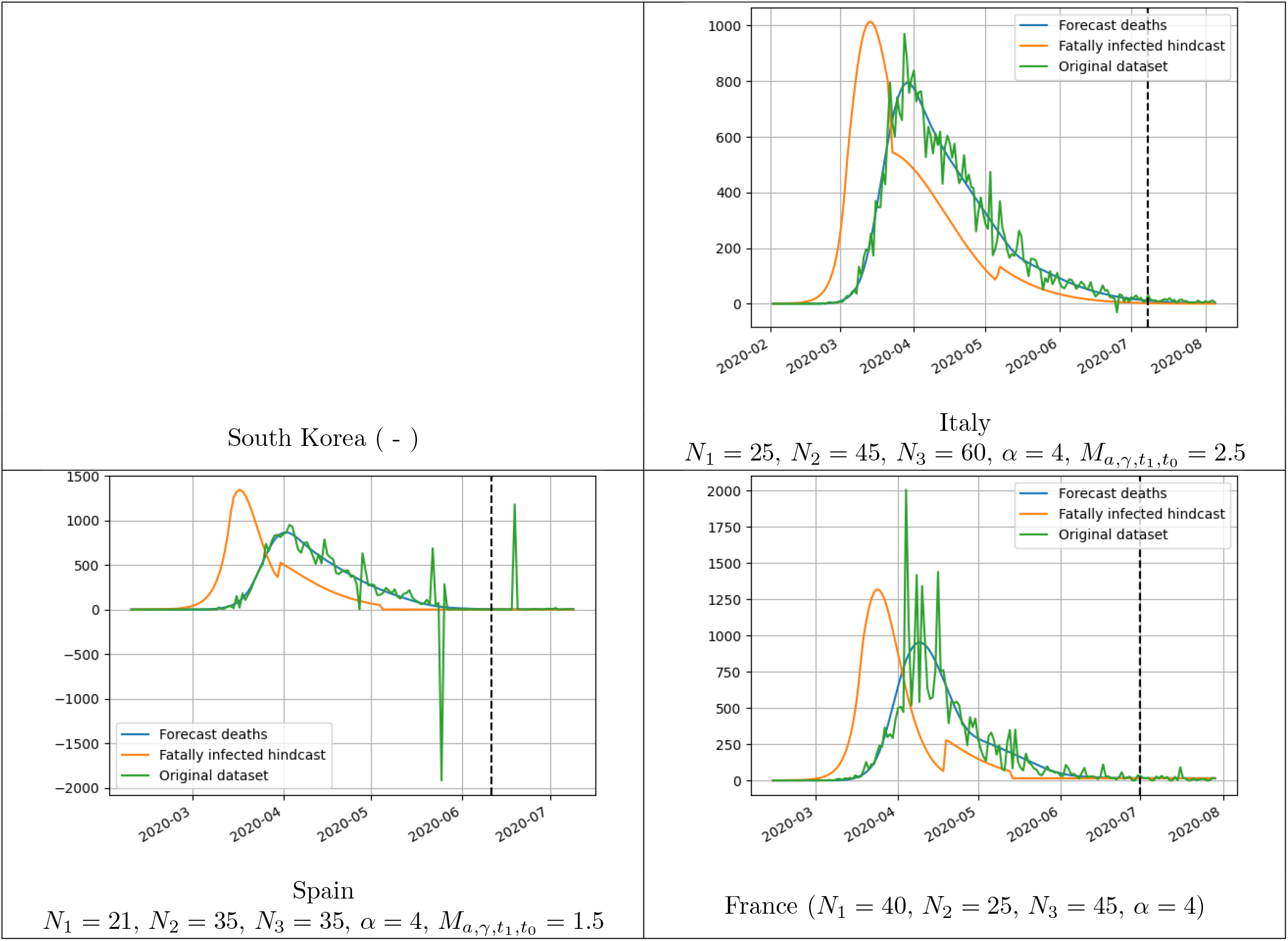
Extended model for deaths applied to Italy, Spain and France. Below the plot of each country we show the parameters of the model manually chosen to get the best fit between the daily registered data and the model prediction. The value of 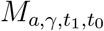(see (5)) only appears if it is manually fixed in the algorithm. We do not include the case of South Korea because the number of deaths were too small to be significant from an statistical point of view.

**Figure 6:**
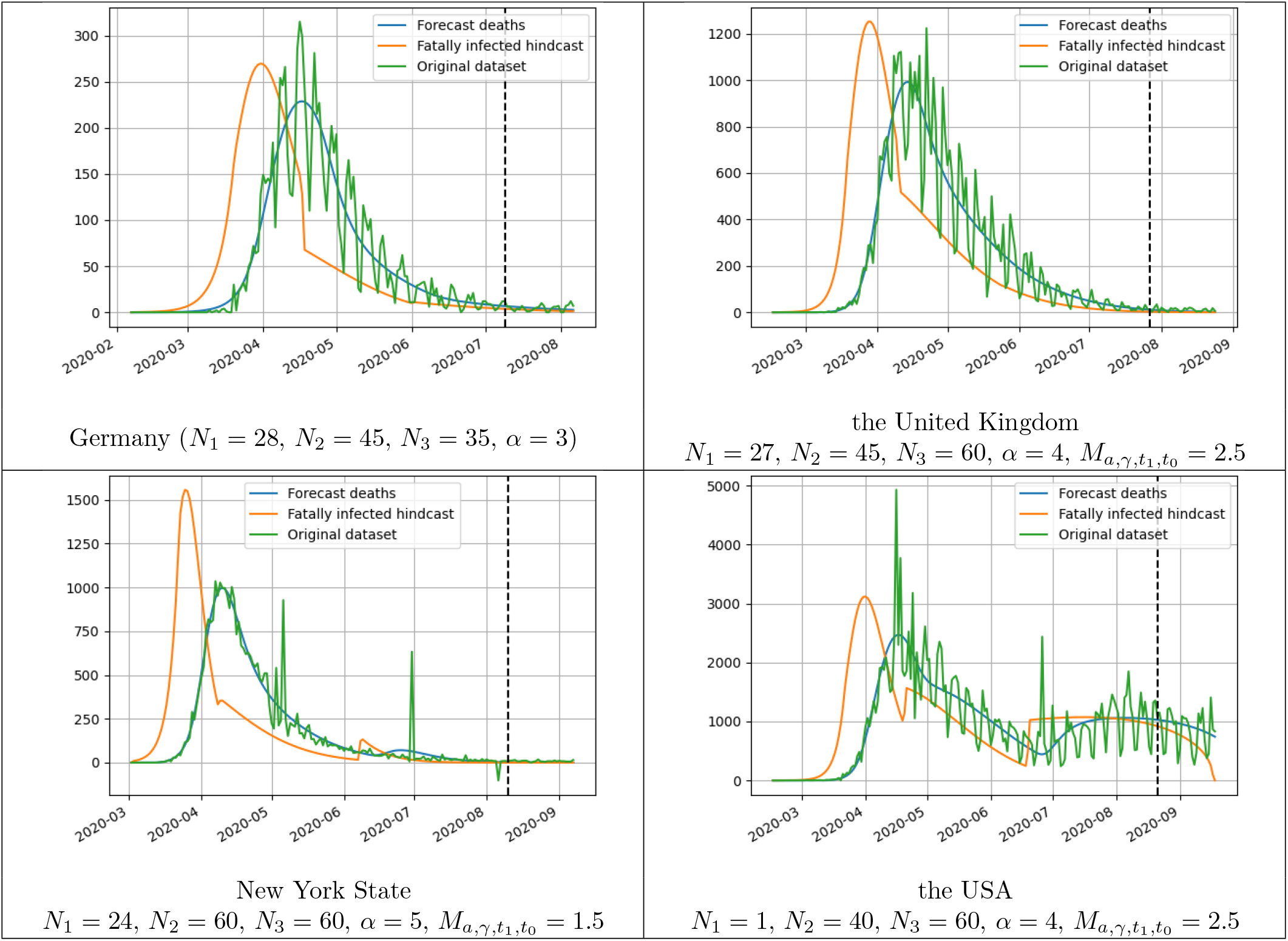
Extended model for deaths applied to Germany, the United Kingdom, New York State and the USA. Below the plot of each country we show the parameters of the model manually chosen to get the best fit between the daily registered data and the model prediction. The value of 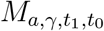(see (5)) only appears if it is manually fixed in the algorithm.

To study the epidemic spread, we will use the timeline (see Fig. 1-2) given by the following epidemiological events:

- **Outbreak**: Start date of the first epidemic wave. In the case of patients who tested positive, we consider that the epidemic wave begins when the accumulated number of observable cases reaches 1 subject per 100,000 inhabitants (1 subject per 1,000,000 in the case of deaths). In fact we have two estimates of the outbreak date. The one obtained using the real data-set communicated by the countries (that we name data outbreak) and the one obtained from the model approximation of the observable data given by *N*(*t*) (that we name model outbreak). In general, there is little difference between the two estimates in the countries studied. In the case of New York state there are 4.4 days of difference in the case of infected due to the fact that very few cases were detected at the beginning of the epidemic.
- **SDM**: Social Distancing Measures. We will assume that the countries implement social distancing measures to control the epidemic. We pay particular attention to cases where a strict lockdown is implemented at the beginning of the epidemic spread because that allows us to study the impact of a strict lockdown as the first measure to control the epidemic. Except in the case of a strict lockdown, this event does not have a specific date associated with it because it can correspond to a variety of measures taken at different times.
- **FTM**: first trend modification in reaction to **SDM**. We assume, in agreement with our model, that at the beginning of the epidemic the coronavirus was in free circulation and that the number of infected grew exponentially with a constant growth rate, *a*_1_, until the accumulated number of infected reached the value 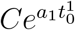 and from that moment, in reaction to the social distancing measures the growth rate began to decrease. We calculate the date when this reaction begins to be observable by the model as the time *t* such that 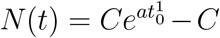. This is a useful information because it tells us the observable reaction time to the social distancing measures. We consider that the coronavirus was in free circulation from the date of the start of the outbreak (computed using the model) until the first trend modification.
- **PEAK**: date when the maximum of *N*′(*t*) is reached in the first wave, which represents the peak of the daily number of observable patients tested positive or deaths.
- **HALF PEAK**: date after reaching the peak when the number of daily cases at the peak is divided by two.
- **EFW**: ending of the first wave. In the case of infected, we observe that over time, the number of new daily cases stabilizes around a baseline that changes between different countries. We manually set the value of the parameter *N*_2_ so that this stabilization time corresponds to *t*_*min*_ + *N*_1_ + *N*_2_. We consider this time as the date of the end of the first wave. In the case of the deceased, this stabilization is not easily observed and we set the date of the end of the first wave as the time the number of new daily cases reaches the value of 1 deceased per 1,000,000 inhabitants.

In tables 2 and 3 we present a summary of the dates of the timeline events we obtain for the different countries for patients tested positive and deaths. Below each date, in brackets, we write the number of days passed since the previous event. Furthermore, for each event we include the value of the daily number of cases predicted by the model for that day (normalized to the country’s population) and below that value, in brackets, we write the ratio between this value and the same value in the previous event. This gives us an idea of the growth rate of the number of daily cases between one event and another. In table 4 we compare between the results obtained for the patients tested positive and deaths. Below we will present a discussion of the most relevant results of these large tables, both globally and country by country.

**Table 1:** Comparison of the estimation of the initial exponential growth rate computed using our method and the method proposed in [7].

|  | S. Korea | Italy | France | Germany | UK |
| --- | --- | --- | --- | --- | --- |
| Method proposed in this paper | 0.2525 | 0.1359 | 0.1282 | 0.2478 | 0.1684 |
| Method proposed in [7] | 0.24 | 0.18 | 0.16 | 0.27 | 0.17 |

**Table 2:** Timeline of the epidemic spread and epidemiological indicators for patients tested positive.

|  |  |  |  |  |  |  |  |  |
| --- | --- | --- | --- | --- | --- | --- | --- | --- |
| **TESTED POSITIVE** | S. Korea | Italy | Spain | France | Germany | UK | New York | the USA |
| **Model Outbreak** | Feb-21 | Feb-29 | Mar-6 | Mar-9 | Mar-9 | Mar-12 | Mar-9 | Mar-14 |
| **Data Outbreak** | Feb-22<br>(+0.666) | Feb-27<br>(-2.04) | Mar-5<br>(-1.02) | Mar-7<br>(-1.92) | Mar-7<br>(-1.11) | Mar-11<br>(-1.23) | Mar-12<br>(+3.3) | Mar-15<br>(+1.27) |
| **Data Outbreak: daily value per 100,000/h** | 0.267 | 0.144 | 0.295 | 0.174 | 0.204 | 0.251 | 1.818 | 0.513 |
| **Lockdown** | - | Mar-11<br>(+12.18) | Mar-14<br>(+8.35) | Mar-17<br>(+9.61) | - | - | - | - |
| **Lockdown: daily value per 100,000/h** |  | 2.89<br>(x 20.05) | 3.51<br>(x 11.91) | 1.7<br>(x 9.75) |  |  |  |  |
| **initial exponential growth rate** | 0.2525 | 0.1359 | 0.1645 | 0.1282 | 0.2478 | 0.1684 | 0.1949 | 0.1869 |
| **FTM: first trend modification** | Feb-29<br>(+6.72) | Mar-18<br>(+7.16) | Mar-22<br>(+8.70) | Mar-26<br>(+9.31) | Mar-17<br>(+9.54) | Mar-25<br>(+14.39) | Mar-25<br>(+12.65) | Mar-27<br>(+12.31) |
| **FTM: daily value per 100,000/h** | 1.15<br>(x 4.31) | 7.181<br>(x 2.49) | 13.58<br>(x 3.86) | 5.27<br>(x 3.10) | 2.062<br>(x 10.11) | 3.31<br>(x 13.18) | 21.34<br>(x 11.74) | 5.03<br>(x 9.81) |
| **PEAK: date** | Mar-1<br>(+1.61) | Mar-25<br>(+7.75) | Mar-27<br>(+5.29) | Apr-2<br>(+6.76) | Apr-1<br>(+14.59) | Apr-9<br>(+15.35) | Apr-8<br>(+14.63) | Apr-8<br>(+11.78) |
| **PEAK: daily value per 100,000/h** | 1.24<br>(x 1.08) | 9.526<br>(x 1.33) | 17.01<br>(x 1.25) | 6.62<br>(x 1.26) | 6.532<br>(x 3.17) | 7.23<br>(x 2.18) | 50.75<br>(x 2.38) | 9.626<br>(x 1.91) |
| **HALF PEAK: date** | Mar-8<br>(+6.37) | Apr-20<br>(+25.09) | Apr-10<br>(+13.08) | Apr-18<br>(+16.04) | Apr-15<br>(+14.70) | May-5<br>(+41.02) | Apr-28<br>(+19.96) | - |
| **EFW: date end first wave** | Apr-25<br>(+47.65) | May-24<br>(+34.00) | May-24<br>(+43.93) | May-12<br>(+23.89) | May-22<br>(+36.23) | Jun-26<br>(+36.00) | Jun-7<br>(+39.25) | - |
| **EFW: daily value per 1,000,000/h** | 0.031<br>(x 0.05) | 0.853<br>(x 0.18) | 0.614<br>(x 0.07) | 1.04<br>(x 0.31) | 0.531<br>(x 0.16) | 0.571<br>(x 0.16) | 3.198<br>(x 0.13) | - |
| **Measure Effectiveness** | 1.0000 | 1.62846 | 1.4927 | 1.4972 | 2.9554 | 2.5 | 2.6237 | 2.2872 |

**Table 3:** Timeline of the epidemic spread and epidemiological indicators for deaths.

| DEATHS | S. Korea | Italy | Spain | France | Germany | UK | New York | the USA |
| --- | --- | --- | --- | --- | --- | --- | --- | --- |
| **Model Outbreak** | - | Mar-3 | Mar-10 | Mar-16 | Mar-18 | Mar-17 | Mar-18 | Mar-23 |
| **Data Outbreak** | - | Mar-3<br>(-0.07) | Mar-11<br>(+1.17) | Mar-13<br>(-3.09) | Mar-22<br>(+4.13) | Mar-17<br>(-0.76) | Mar-18<br>(+0.09) | Mar-21<br>(-1.31) |
| **Data Outbreak:** daily value per 1.000.000/h |  | 0.29 | 0.513 | 0.11 | 0.312 | 0.24 | 0.446 | 0.188 |
| **Lockdown** | - | Mar-11<br>(+7.70) | Mar-14<br>(+2.02) | Mar-17<br>(+3.76) | - | - | - | - |
| **Lockdown:** daily value per 1.000.000/h |  | 2.06<br>(x 7.19) | 0.92<br>(x 1.80) | 0.38<br>(x 3.50) |  |  |  |  |
| initial exponential growth rate | - | 0.2498 | 0.2517 | 0.2486 | 0.1673 | 0.2497 | 0.2560 | 0.2498 |
| **FTM:** first trend modification | - | Mar-16<br>(+5.94) | Mar-27<br>(+13.76) | Mar-31<br>(+14.52) | Apr-02<br>(+11.35) | Mar-31<br>(+14.61) | Apr-5<br>(+18.87) | Apr-4<br>(+13.10) |
| **FTM:** daily value per 1,000,000/h | - | 6.173<br>(x 2.99) | 15.44<br>(x 16.72) | 9.45<br>(x 24.54) | 1.56<br>(x 4.98) | 6.83<br>(x 28.45) | 42.70<br>(x 95.74) | 3.49<br>(x 18.54) |
| **PEAK:** date | - | Mar-29<br>(+12.36) | Apr-1<br>(+5.06) | Apr-9<br>(+8.58) | Apr-16<br>(+13.83) | Apr-13<br>(+12.03) | Apr-10<br>(+5.03) | Apr-17<br>(+13.08) |
| **PEAK:** daily value per 1,000,000/h | - | 13.142<br>(x 2.13) | 18.52<br>(x 1.20) | 14.58<br>(x 1.54) | 2.73<br>(x 1.76) | 14.62<br>(x 2.14) | 51.325<br>(x 1.20) | 7.459<br>(x 2.14) |
| **HALF PEAK:** date | - | Apr-25<br>(+27.11) | Apr-21<br>(+19.77) | Apr-23<br>(+13.97) | May-4<br>(+17.30) | May-4<br>(+20.79) | Apr-25<br>(+14.91) | - |
| **EFW:** ending first wave | - | Jun-9<br>(+45.25) | May-21<br>(29.96) | May-28<br>(+35.80) | May-9<br>(+5.22) | June 24<br>(+50.73) | Jul-18<br>(+83.69) | - |
| Measure Effectiveness | - | 2.5 | 1.5 | 1.9297 | 2.1677 | 2.5 | 1.5 | 2.5 |

**Table 4:** Comparison between the spread of patients tested positive and the spread of of deaths in different countries. We compare the lag between the start of the outbreaks, the lag between the peaks and the fatality rate computed as the ratio between the values in the peaks.

|  | S. Korea | Italy | Spain | France | Germany | UK | New York | the USA |
| --- | --- | --- | --- | --- | --- | --- | --- | --- |
| Lag between data outbreaks | - | 4.48 | 6.33 | 5.85 | 14.68 | 5.93 | 5.59 | 6.45 |
| Lag between peaks | - | 3.39 | 4.83 | 7.03 | 15.73 | 3.83 | 2.22 | 8.53 |
| Fatality rate ratio peak values | - | 13.80% | 10.89% | 22.03% | 4.18% | 20.17% | 10.11% | 7.75% |

### 3.0.1 Analysis country by country

#### South Korea

Since the initial outbreak, the virus was in free circulation only for 7.38 days, then the model starts to react to the **SDM** and reached the daily peak 1.61 days later and the number of cases at the peak (1.24) was divided by two 6.37 days later. A very good performance in controlling the epidemic spread. South Korea monitored the spread of the epidemic using technological resources, such as tracking credit card use, checking CCTV footage and an efficient centralized contact-tracing using cellular phones. The number of deaths has been very small and has not been studied because it is not statistically significant.

#### Italy

On 8 March 2020, a strict lockdown was imposed in the Lombardy region and later, on March 11, the lockdown was extended to the whole country. Since the initial outbreak, the virus was in free circulation for 17.31 days (7.16 days after the second lockdown). The peak of daily cases was reached 7.75 days later and the number of cases at the peak (9.526) was divided by two 25.09 days later. The fact that the lockdown in Lombardy was implemented 3 days before the lockdown in the whole country modifies the reaction time with respect to the second lockdown.

#### Spain

On 14 March 2020, a strict lockdown was imposed in the whole country. Since the initial outbreak, the virus was in free circulation for 16.03 days (8.7 days after the lockdown). The peak of daily cases was reached 5.29 days later and the number of cases at the peak (17.01) was divided by two 13.08 days later. The first really significant measure to control the epidemic that Spain and France implemented was the lockdown. Therefore, these two countries are a good example to study how a strict lockdown affects the free circulation of the virus.

#### France

On 17 March 2020, a strict lockdown was imposed in the whole country. Since the initial outbreak, the virus was in free circulation for 17 days (9.31 days after the lockdown). The peak of daily cases was reached 6.76 days later and the number of cases at the peak (6.62) was divided by two 16.04 days later.

#### Germany

Since the initial outbreak, the virus was in free circulation for only 8.43 days. The peak of daily cases was reached 14.59 days later and the number of cases at the peak (6.532) was divided by two 14.70 days later. A good performance in controlling the epidemic spread.

#### the United Kingdom

Since the initial outbreak, the virus was in free circulation for 13.16 days. The peak of daily cases was reached 15.35 days later and the number of cases at the peak (7.23) was divided by two 41 days later.

#### New York State

Since the initial outbreak, the virus was in free circulation for 15.94 days. The peak of daily cases was reached 14.63 days later and the number of cases at the peak (50.75) was divided by two 19.96 days later.

#### the USA

Since the initial outbreak, the virus was in free circulation for 13.59 days. The peak of daily cases was reached 11.78 days later. In the USA each state followed different strategies at different times, globally we cannot say that a first wave has been completed in the USA and then, in this case, we do not study the evolution in the USA after the first peak.

### 3.0.2 Global Analysis

At the beginning of the epidemic spread, among the different countries studied, only South Korea and Germany had a testing capacity that allowed to correctly follow the evolution of the epidemic. Indeed, these two countries obtain a similar and high initial exponential growth rate (around 0.25) that can be obtained because of a good testing capacity which is able to track the epidemic spread. Furthermore, in the case of Germany there are other indicators that suggest this fact, such as the high delay in the number of days between the outbreak of infected and the outbreak of deaths and between the dates at the peaks of daily cases of infected and deaths. The mortality ratio obtained by Germany is also a good sign of the testing capacity. An important key to the success of these two countries has been the anticipation reflected in the small number of days the virus circulated freely before social distancing measures began to take effect.

In the rest of the countries, the testing capacity was not good enough to follow the evolution of the epidemic during the first wave. In short, the numbers of infected was strongly underestimated. In these countries the initial exponential growth rate is lower than 0.2, there is little delay between the outbreaks of infected and deaths and the peaks and in all these countries the mortality ratio is artificially high.

In [7] the authors computed an initial exponential growth rate, (*a*_1_ following our notation), for different countries. In table 1 we show some comparison results. The results are reasonably similar considering that the techniques are quite different. In [7] the calculation was carried out directly on the registered data of infected and we do it on the infected hindcast (notice that we always estimate the infected hindcast) and in [7], the authors use a time interval set manually and we use a time interval calculated automatically when minimizing the quadratic error.

In Italy, Spain and France, who implemented a lockdown in the early stages of the epidemic spread, it took between 8 and 10 days to begin to notice the effect of this measure (a little earlier in Italy due to the lockdown in Lombardy). This is quite a reasonable delay considering the incubation time and the administrative time required to register the cases. Afterwards, it took between 5 and 7 days to reach the peak of daily cases (a little longer in Italy due to the Lombardy effect again). In Germany, UK and New York it took between 14 and 16 days to reach the peak of daily cases since the effect of **SDM** began to be noticed. This suggests that the lockdown accelerated this phase considerably. The later time to divide by two the number of daily cases reached at the peak is highly variable among all countries and it cannot be clearly concluded that the initial lockdown improves results.

The social distance effectiveness measure given by 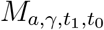 is very good in South Korea 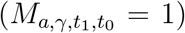. In countries who implemented a strict lockdown it is about 1.5 (it is a little higher in Italy because of the early Lombardy lockdown) and in the other regions it is higher than 2. Even in Germany the value is quite high (2.9554). The reason is that this measure of effectiveness considers the evolution of the epidemic spread after the **SDM** starts to be noticed. So in the case of Germany the **SDM** starts to be noticed very soon but the effect of the **SDM** was not so clear as in the case of a strict lockdown.

In the study of the evolution of the number of deaths, it is highlighted that the initial exponential growth rate of those infected who later die is very similar among all countries except Germany. The reason could be that the number of deaths is independent of the testing capacity, but the calculation of the initial exponential growth rate requires, for the deaths, that the virus has been circulating freely for a sufficient number of days, which happens in all countries except Germany and this affects the evolution of the number of deaths. We observe that the initial exponential growth rate for those infected who later died estimated in Italy, Spain, France, UK, New York State and the USA are very similar to each other and in turn very similar to the growth rate of those infected who tested positive in South Korea and Germany. The mean of all these values obtained in these countries is 0.2507375 with a standard deviation of only 0.0024433264. This seems to indicate that the initial exponential growth rate when the virus circulates freely is about 0.2507375. One of the key parameters in the initial epidemic dynamics is the doubling time *t*_*D*_ which according with our estimation corresponds to

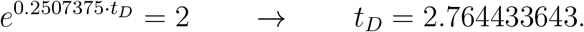

This estimate is consistent with the result shown in [5] where the authors estimate that *t*_*D*_ ∈ [1.86, 2.96]. Another key parameter of the initial epidemic dynamics is the reproduction number *R*0. There are a variety of techniques to compute *R*0 from the initial exponential growth rate, *a*_1_, and some other epidemiological information like the serial time or the mean recovery time 1*/γ*. If we assume that the mean recovery time is 1*/γ* ∈ [7, 14] then we have that

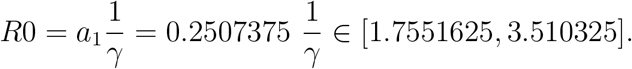

An evident key to success or failure to control the epidemic in the first wave was the anticipation in decision-making that is reflected in the number of days that the virus had been circulating freely before the social distancing measures began to take effect. According to our calculations, each day that the virus circulated freely, the number of infected was multiplied by a factor of 1.285 = *e*^0.2507375^. This indicates the enormous importance of anticipation when taking social distancing measures.

## 4 The ability of the exponential model to predict the epidemic evolution in advance

To study the ability of the model to predict the evolution of the epidemic in the early stages of the epidemic spread we focus on the study of the model estimation, using the available data up to a given date, of the following epidemiological events:

- The date the peak of new daily cases is reached.
- The value of the number of daily cases at the peak.
- The 7-day average of the accumulated number of cases three weeks after reaching the peak.

We are going to use the case of France in this study. The case of France is quite challenging because both the evolution of the number of registered infected and the evolution of the number of deaths show strong fluctuations near the peak of new daily cases. The parameter *N*_1_ indicates the number of days used to compute the model parameters. Therefore, modifying *N*_1_ we obtain the model estimation using the data up to a given date which depends on the value of *N*_1_. In Fig. 7 and 9 we illustrate the basic model for the number of patients tested positive and deaths and some particular values of *N*_1_. We will compare the results obtained by leaving free the effectiveness of the social distancing measures given by 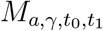 and setting “a priori” the value of 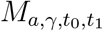. In all the experiments presented in this section we use as regularization weight parameter *α* = 0.

**Figure 7:**
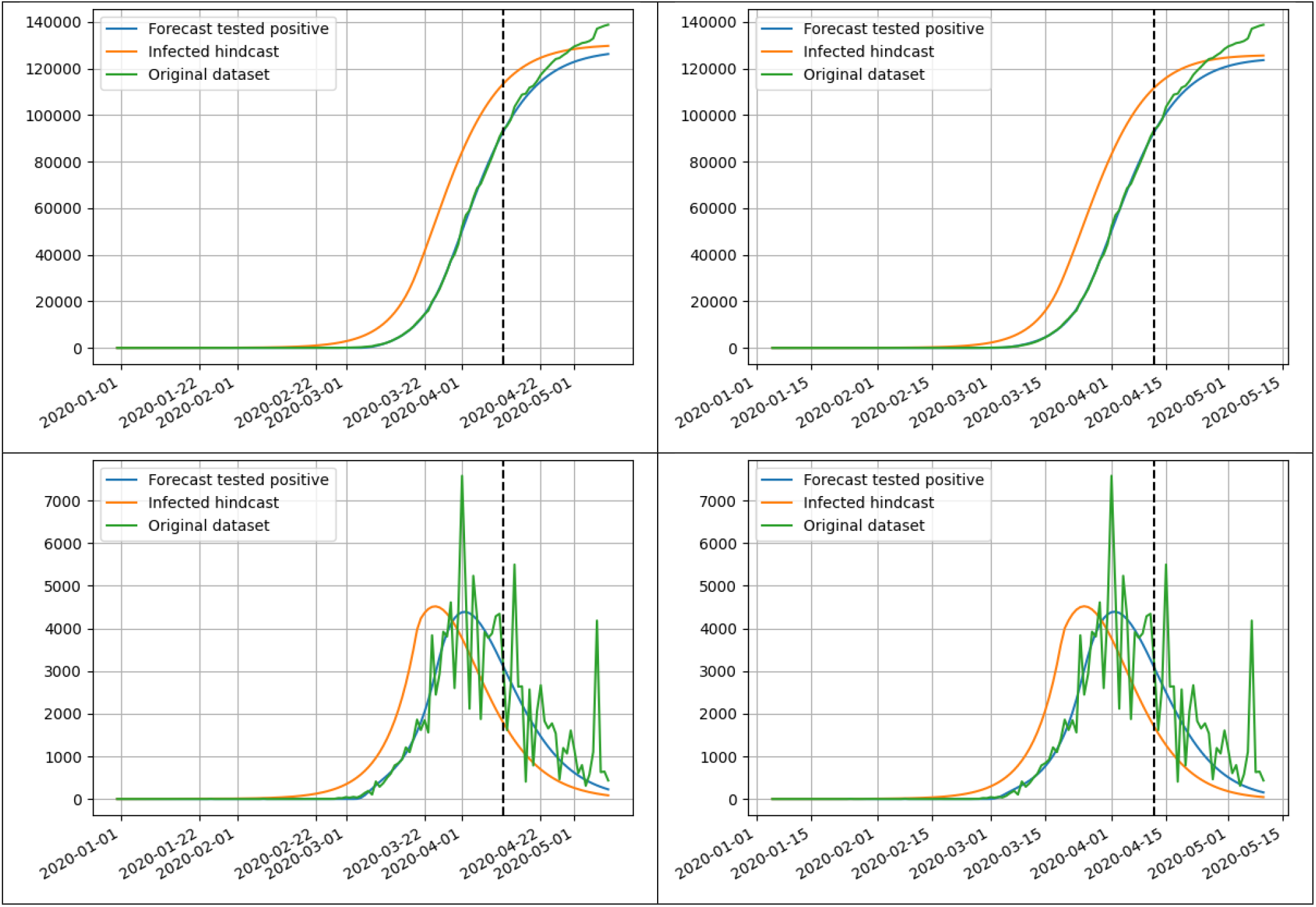
Basic model for patients tested positive in France using *N*_1_ = 45. On the left we present the accumulated and daily cases when the effectiveness is free and on the right when the effectiveness is fixed to 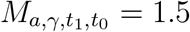. The algorithm uses the data up to the date represented by the vertical black line.

**Figure 8:**
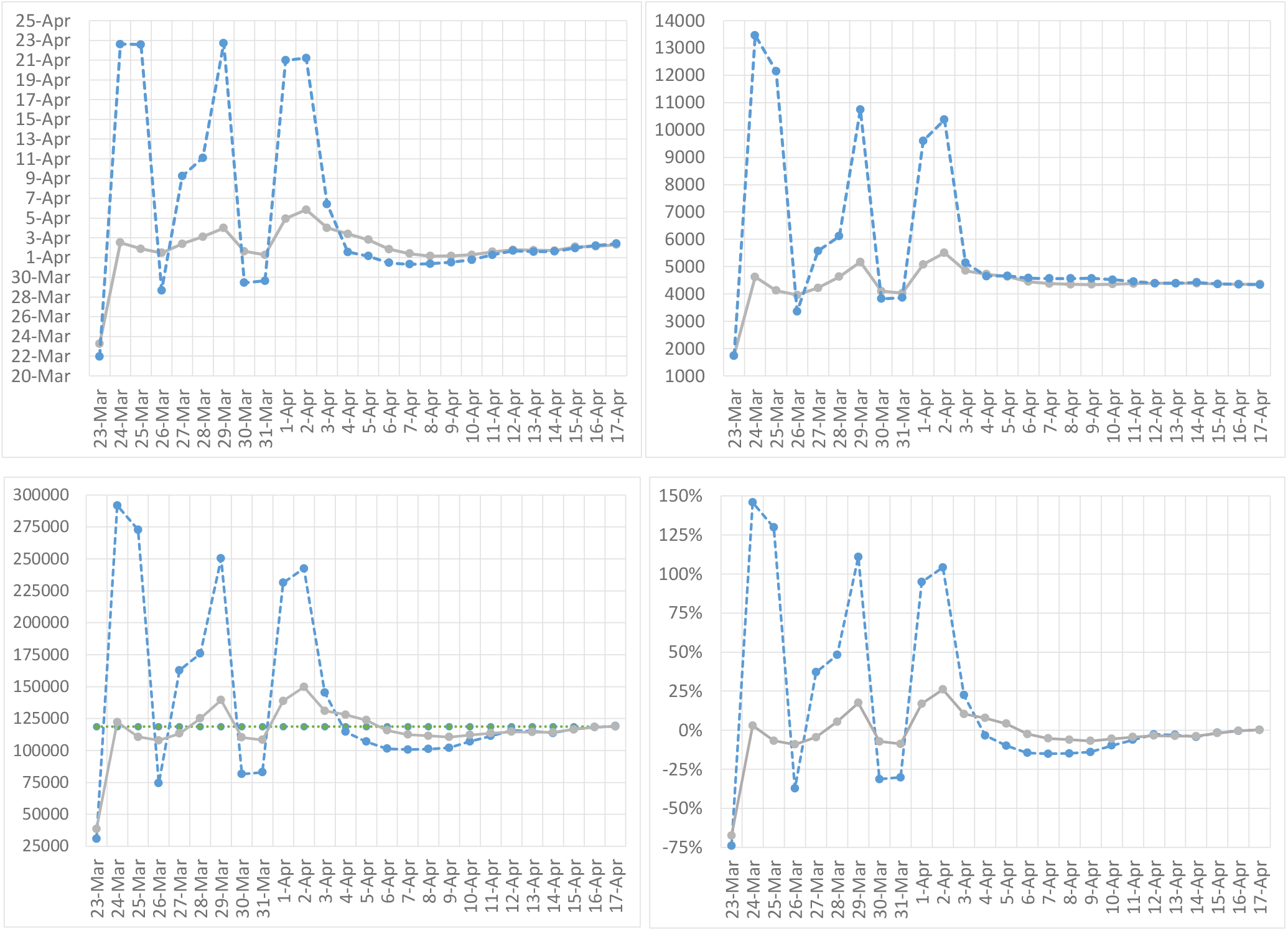
Some epidemiological indicators computed by our model using the registered number of new infected up to the date in the horizontal axis. The solid lines represent the estimate constraining the effectiveness 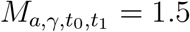, the dashed line, the estimate without constraining 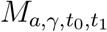, and the dotted line represents the average of the cumulative registered number of infected between April 20 and April 26. From left to right and from top to down, we present: (i) the estimated date of peak of the daily number of patients tested positive, (ii) the estimated value of the daily number of patients tested positive in the peak, (iii) the estimate of the average of the cumulative number of patients tested positive between April 20 and April 26, and (iv) the error, in percentage, between the estimate and actual value of the average of the cumulative number of patients tested positive between April 20 and April 26.

**Figure 9:**
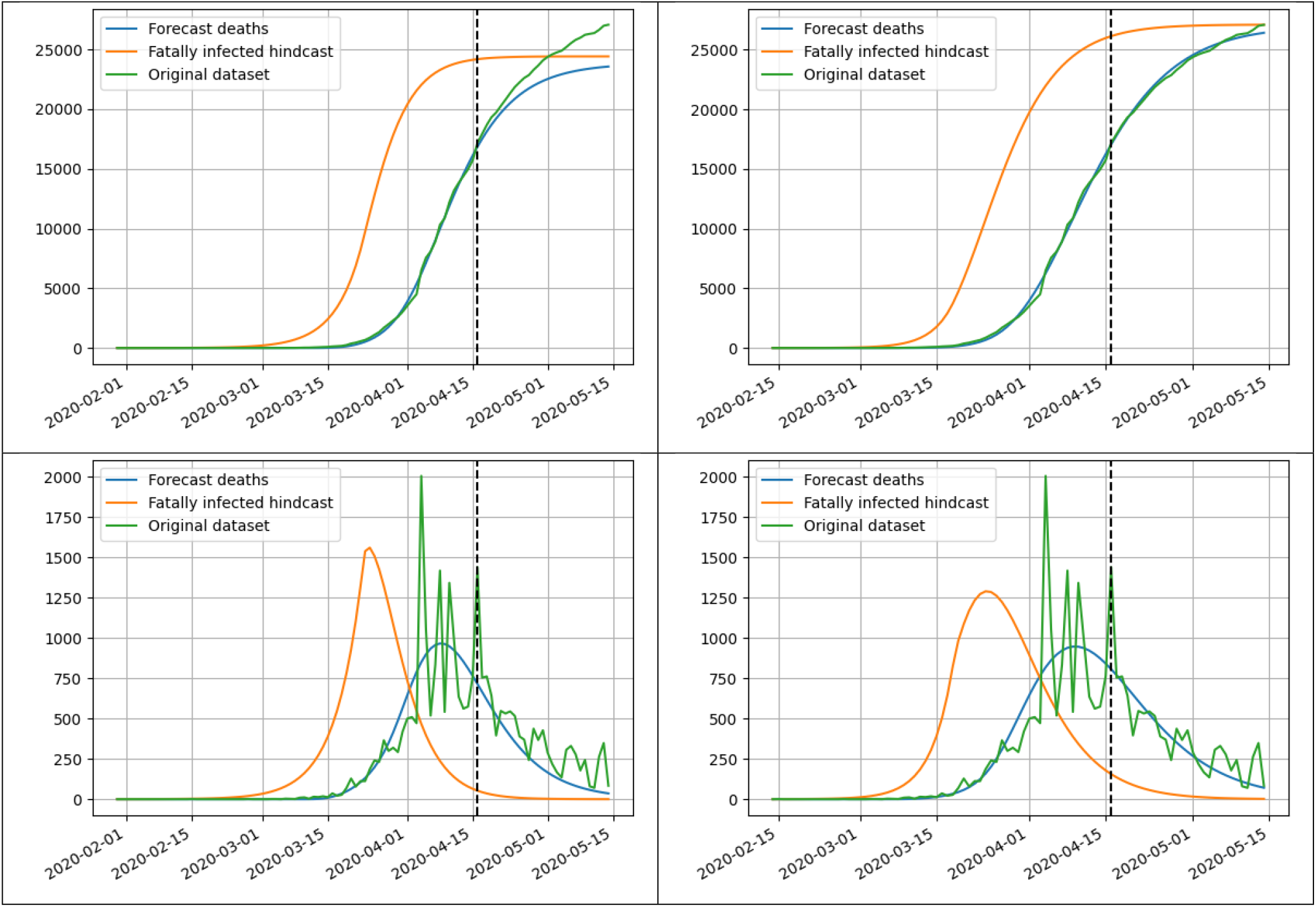
Basic model for deaths in France using *N*_1_ = 34. On the left we present the accumulated and daily cases when the effectiveness is free and on the right when the effectiveness is fixed to 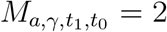. The algorithm uses the data up to the date represented by the vertical black line.

In Fig.8 and 10 we show the estimate of the epidemiological events explained above using the available data up to a given date. We point out that a strict lockdown was implemented in France by March 17, the peak of the daily new cases was reached at April 2 (for patients tested positive) and in April 9 (for deaths). In the case of patients tested positive, we observe that, without constraining the value of 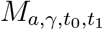, we obtain a reasonable estimate of the epidemiological events since April 3 (that is 1 day after reaching the peak). However, if we fix 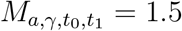, we obtain a reasonable estimate since March 24, just 7 days after the lockdown implementation and 9 days before reaching the peak, which means that the model was able to predict the events quite correctly in advance. In fact, the error in the estimation of March 24 was quite small (just 2.92% in the estimation of the 7-day average 3 weeks after the peak).

**Figure 10:**
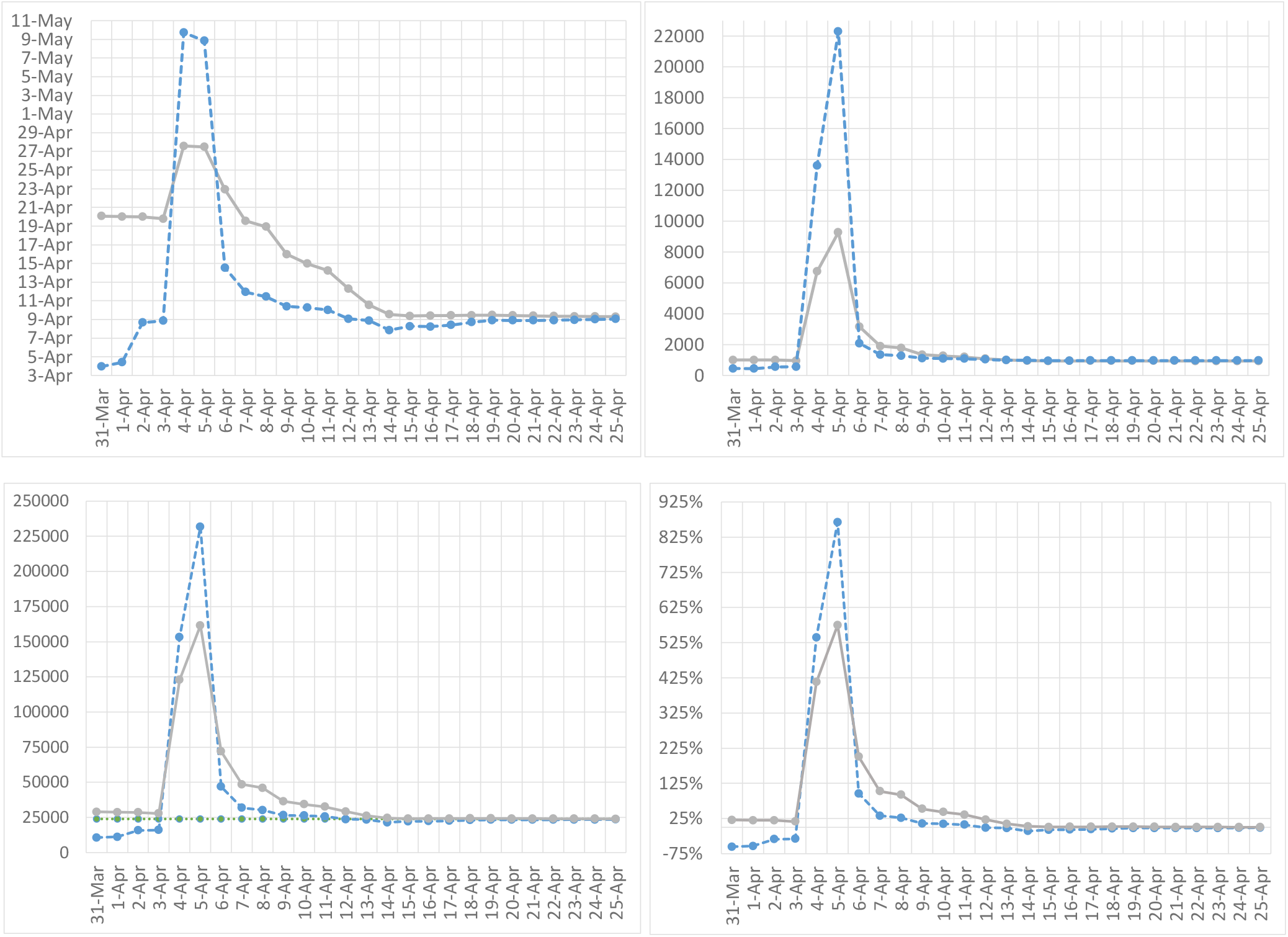
Some epidemiological indicators computed by our model using the registered number of deaths up to the date in the horizontal axis. The solid lines represent the estimate constraining the effectiveness 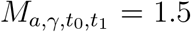, the dashed line, the estimate without constraining 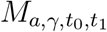, and the dotted line represents the average of the cumulative registered number of deaths between April 27 and May 3. From left to right and from top to down, we present: (i) the estimated date of the daily peak of the new deaths, (ii) the estimated value of the daily number of deaths in the peak, (iii) the estimate of the average of the cumulative registered number of deaths between April 27 and May 3, and (iv) the error, in percentage, between the estimate and actual value of the average of the cumulative registered number of deaths between April 27 and May 3.

In the case of deaths, the results are not so good. On April 4, a large number of new deaths were recorded in France, which produced a great disturbance in the results of the model. In fact, the estimate of April 3 is much better than that of April 4, and until April 7 (two days before the peak) it does not begin to provide a reasonable estimate of the events analyzed. In this case setting the value of 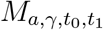 does not contribute much and in fact a correct approximation is obtained before using the model without setting the value of 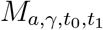.

## 5 Conclusions

Using an exponential model with time-varying growth rate we study in details the chronology of the main epidemiological events during the full course of the first wave in South Korea, Italy, Spain, France, Germany, the United Kingdom, the USA and the New-York state. This model allows us to estimate interesting features as the reaction time of the observed registered cases to strict social distancing measures, or the time the coronavirus has been in free circulation at the beginning of the epidemic. Comparing the results of the different regions we can conclude that a good testing capacity and anticipation in implementing social distancing measures has been very important in controlling the epidemic during the first wave.

Using the obtained results, we can observe the lack of an adequate testing capacity in several ways: (i) a lower than expected estimate of the exponential growth rate when the coronavirus is in free circulation, which means that the testing capacity is not able to track the actual growth of the epidemic, (ii) a short delay between the dates detected for the number of patients tested positive and deaths of events such as the epidemic outbreak or the peak of daily cases, or (iii) a high mortality rate measured as the ratio between the values of the peaks of daily death cases and positive tests.

Surprisingly, we obtain, when the virus is in free circulation, that eight values of the exponential growth rate obtained from the infected hindcast of South Korea and Germany for the patients tested positive and for Italy, Spain, France, UK, the USA and the New York State for deaths are very similar to each other with a mean of 0.2507375 and a standard deviation of only 0.0024433264. This standard deviation is very small and suggests that the value 0.2507375 is a good estimate of this exponential growth rate.

We have also studied the ability of the model to properly forecast the epidemic spread at the beginning of the epidemic outbreak when very little data and information about the coronavirus were available. In the case of a strict lockdown implementation and constraining the value of the effectiveness 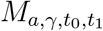of the lockdown (based on the results obtained for other countries), we obtain, in the case of France a reasonable estimate of the peak of the new cases of patients tested positive 9 days in advance and only 7 days after the implementation of the lockdown. We believe that the success of this prediction is based on the simplicity of the model and its low number of parameters, which compensates, to some extent, for the low quality of the data used.

All the experiments we have presented can be reproduced using the online interface www.ctim.es/covid19. For more technical information about the used exponential model see [1].

## Data Availability

All data referred to in the manuscript are available at the European Centre for Disease Prevention and Control

https://www.ecdc.europa.eu/en

